# Early risk factors for metabolic dysfunction in young people with major mood disorders: Longitudinal path analysis of a prospective birth cohort using structural equation modelling

**DOI:** 10.1101/2025.08.24.25334329

**Authors:** S. McKenna, M. Varidel, M. Shin, J. Carpenter, E. Tonini, J. Crouse, E.S. Scott, A. Mamun, J.G. Scott, J. Najman, I.B. Hickie

**Affiliations:** Brain and Mind Center, The University of Sydney, NSW, Australia; UQ Poche Centre for Indigenous Health, The University of Queensland, Brisbane, Australia; ARC Centre of Excellence for Children and Families over the Life Course, The University of Queensland, Brisbane, Australia; Child and Youth Mental Health Service, Children’s Health Queensland, QLD, Australia; Australia Child Health Research Centre, The University of Queensland, QLD, Australia; School of Public Health, The University of Queensland, QLD, Australia; School of Social Sciences, The University of Queensland, QLD, Australia

## Abstract

**Objectives:** Lifestyle interventions targeting weight loss currently show limited benefits for individuals with major mood disorders. Sleep-wake disturbances are prevalent in youth with mood disorders and linked to metabolic dysfunction. This study evaluates whether sleep-wake disturbance is a driver of metabolic dysfunction for individuals with major mood disorders, as compared to the existing theory that weight gain is the main contributor.

**Design:** Longitudinal cohort study in 1712 individuals aged 21 to 30 years. Spearmans’ correlation coefficients between depressive symptoms, sleep-wake cycle disturbance, and metabolic variables at 21 and 30 years. Structural equation modelling explored: 1. A cross-lagged effects model of depressive symptoms, sleep-wake cycle disturbance and body mass index (BMI), and 2. A parallel mediation model comparing two indirect pathways between depressive symptoms at 21 years and HOMA2-IR (a measure of insulin-glucose homeostasis) at 30 years through sleep-wake disturbance and BMI.

**Results:** We found a small correlation between depressive symptoms at 21 and HOMA2-IR at 30 (*r*=0.07, p<0.01). In the mediation model, the direct relationship between depressive symptoms at 21 and HOMA2-IR at 30 was no longer significant but there was a small total relationship (*B=*0.05, *p*<0.05) and a significant indirect pathway through sleep-wake disturbance (*B=*0.01, *p*<0.05). Although not a significant mediator, BMI at 30 was strongly related to HOMA2-IR (*B=*0.44, *p* <0.001).

**Conclusion:** In this study, BMI was not a key mediator of the relationship between depressive symptoms and HOMA2-IR, while sleep-wake cycle disturbance appeared to play a small role. Work is needed in clinical populations with more robust measures of sleep-wake disturbance.

---

Individuals with major mood disorders have 15 to 20 year reduced life expectancy^1–3^ and the mortality gap has not meaningfully improved in the last decade^4,5^, suggesting novel intervention approaches are urgently needed. A major contributor is increased rates of premature cardiovascular disease (CVD), largely attributable to the prevalence of metabolic syndrome in this population.^6,7^ Weight gain and smoking are assumed to be key modifiable risk factors however there is increasing evidence that more complex pathophysiological processes are involved. For example, sleep-wake cycle disturbances, such as those evident in shift workers, are associated with metabolic dysfunction including increased insulin resistance, plasma glucose, and triglycerides.^8,9^ Sleep-wake disturbances are also common in youth with major mood disorders and associated with more severe disorders.^10,11^ However, it is unclear whether sleep-wake disturbances co-occur or are causally related to metabolic dysfunction in individuals with major mood disorders.

Young people with mood disorders present a unique opportunity to examine potential mechanisms of metabolic dysfunction. At this early stage of illness, they are likely to have had minimal exposure to confounding factors such as long-term medication use and chronic weight gain that typically occur later in the disorder trajectory. Despite this relatively lower risk profile for CVD and rates comparable to the general population of being overweight/obese, and normal glucose levels^12–14^, young people presenting to early intervention mental health services show elevated HOMA2-IR (Homeostatic Model Assessment for Insulin Resistance) levels Early work has shown that a sub-group of young people with mood disorders and sleep-wake disturbances have significantly higher HOMA2-IR levels as compared to other mood disorders, but not higher body mass index (BMI).^15^ Moreover, sleep interventions such as CBT-I (Cognitive Behavioural Therapy for Insomnia) and melatonin have been shown to improve HOMA2-IR in youth populations with major mood disorders.^16–18^

The current study utilised a large prospective youth cohort (21 to 30 years), taken from the Mater-University Study of Pregnancy (MUSP), to test a novel hypothesis, that sleep-wake cycle disturbance is a major driver of abnormal glucose-insulin homeostasis early in the course of mood disorders, as compared to the existing assumption that it is driven solely by weight gain (for instance due to unhealthy lifestyle factors and medication use). If correct, there would be increased impetus for detailed longitudinal tracking of sleep-wake cycle disturbance and metabolic blood markers (particularly HOMA2-IR) in youth cohorts with major mood disorders, and a need to test novel intervention and prevention strategies, to reduce CVD risk and improve the mortality gap.

## METHODS

We used data from a prospective birth cohort study of mothers and their offspring in MUSP. Participants were the offspring of pregnant women who had delivered a singleton baby at a major public hospital in South Brisbane between 1981 and 1983. Written informed consent from mothers was obtained at all data collection phases and from offspring at 21-and 30-years follow-up (hereafter ‘years’ rather than ‘years follow-up’ for simplicity). Baseline data were collected from 7,223 mothers and their offspring and follow-up data was collected at 6-months, and 5, 14, 21, and 30 years. The 21-year data collection was conducted in 2002, the 30-year was conducted between 2011 and 2014. We included 1712 children (62% female; see Table 1) who had completed questionnaires and physical assessments at 21 years and who had provided questionnaires, physical assessments, and blood chemical data at 30 years. Ethical approval for this study was obtained from The University of Queensland Human Research Ethics Committee (mothers: B/555/SS/01/NHMRC - 30/11/2001, offspring: B/660/SS/01/NHMRC - 20/12/2001) and The Mater Human Research Ethics Committee (mothers: 505A - 29/6/2002, offspring: 506A - 15/07/2002).

**Table 1.** Descriptive statistics of demographic variables, depression, sleep-wake disturbance, and metabolic outcomes.

| VARIABLE | 21 Years old |  |  | 30 Years old |  |  |
| --- | --- | --- | --- | --- | --- | --- |
|  | M(SD) | Range | N | M(SD) | Range | N |
| Age |  |  |  | 30.24 | 27.81 – 32.94 | 1,712 |
| Gender (% Female) |  |  |  | 62% |  | 1,712 |
| Depression (CESD) | 11.00<br>(8.71) | 0 - 60 | 1,517 | 10.02<br>(9.40) | 0 - 60 | 1,599 |
| Sleep-wake disturbance (PSQI)<sup>1</sup> | 2.49<br>(1.56) | 0 - 12 | 1,522 | 5.43<br>(3.16) | 0 - 21 | 1,512 |
| Metabolic variables |  |  |  |  |  |  |
| *BMI* | 24.37<br>(5.05) | 14.48 - 51.64 | 1,133 | 27.14<br>(6.15) | 15.65 – 67.74 | 1,344 |
| *HOMA2_IR* |  |  |  | 1.14<br>(1.09) | 0.16 – 16.13 | 1,683 |

<sup>1</sup>A shortened version of the PSQI was used at 21 years old.**Table 2.**

### Measures

#### Depressive symptoms

Depressive symptoms were assessed at 21 and 30 years using the Center for Epidemiologic Studies Depression Scale (CES-D).^19^ The CESD is a 20-item measure of depressive symptoms in the previous week. Participants are asked to rate items such as “I felt depressed” and “I felt that people disliked me” on a 0 to 3 scale from “rarely or none of the time (less than 1 day)” to “most or all of the time (5-7 days)”. This scale has been shown to have high internal consistency, acceptable test-retest stability, and strong concurrent and construct validity based on alternative clinician and self-rated measures.^20^ Scores above 16 have been recommended as the cutoff score to identify depression.^19^ Depressive symptoms were measured identically at 21 and 30 years.

#### Sleep-wake disturbance in young adults

In this study, sleep-wake disturbance was assessed by the Pittsburgh Sleep Quality Index (PSQI).^21^ The PSQI is a comprehensive, self-report questionnaire that involves seven clinically relevant domains of sleep including subjective sleep quality, sleep latency, sleep duration, habitual sleep efficiency, sleep problems (i.e. snoring, too cold), use of sleep medication, and daytime dysfunction. As reported previously^22^, at 21 years only a shortened version of the PSQI was available (5/7 domains were available including subjective sleep quality, sleep duration, sleep medication, sleep disturbance, and daytime dysfunction). All components were available at 30 years. Each domain is an ordinal variable that is coded from 0 – 3 where 0 indicates “no difficulty” and 3 “severe difficulty” (participants were asked to report sleep habits during the past month only). Scores from these separate components were combined to derive a global measure of sleep disturbance. The PSQI has been shown to have strong psychometric properties in youth populations and in populations with mood disorders.^23,24^

#### BMI and cardiometabolic measures

Physical assessments were conducted at both 21 and 30 years. BMI (weight (kg)/height^2^) was obtained by measuring height using a portable stadiometer and weight using the Tanita Body Composition Analyser BC418. Blood chemical data were available only at 30 years. Insulin, glucose and blood lipids were measured in a fasting blood sample. Respondents were asked to eat by 7 pm the previous evening and fast for at least 9 h before blood sampling. Samples were collected by Mater Pathology Service, Brisbane. Respondents who lived outside the Brisbane area had their samples obtained by a participating laboratory. The homeostatic model assessment of insulin resistance (HOMA2-IR) was estimated using fasting insulin (m u/L) and fasting glucose (mmL/L).

#### Covariates

We adjusted for participants’ gender and age during all analyses. We also adjusted for physical activity and smoking at age 30. Participants rated their activity using the International Physical Activity Questionnaire (IPAQ)^25^, a 7-item questionnaire with good psychometric properties^25^, consisting of open-ended questions surrounding the individuals’ activity levels over the past week. For example, “during the last 7 days on how many days did you do vigorous physical activities like heavy lifting, digging, aerobics, or fast bicycling?”. Participants self-reported their frequency of smoking and were given a score from 0 (0 cigarettes per day) to 3 (20+ per day).

### Data analysis

All analysis were conducted in R (v. 0.6.19).^26^ P-values of less than 0.05 on both the Kolmogorov–Smirnov and Shapiro– Wilk tests along with visual inspection of variables indicated that the data did not have a normal distribution. Variables were centred by subtracting the mean and then dividing by the standard deviation, creating a new variable with a mean of 0 and a standard deviation of 1. Missing data were handled through multivariate imputation by chained equation using the MICE package^27^ to maximise data available for analysis (see Table S1 for a summary of missing data [N and %]).

Direct bivariate relationships between depressive symptoms, sleep and cardiometabolic variables were explored, while controlling for age and gender, using Spearman’s correlation coefficient. This method is robust when exploring non-normally distributed data. Path analysis (controlling for covariates) was conducted in R using the lavaan package (v. 0.6.19).^26^ To test the fit of the models, maximum likelihood estimation was employed, using the following fit indices: the chi-square (χ^2^) statistic, the comparative fit index (CFI), the Tucker-Lewis index (TLI), the root mean square error of approximation (RMSEA), and the standardized root mean residual (SRMR). If χ^2^ is not significant, the CFI and TLI values are.90 or higher, and the SRMR and RMSEA values are.10 or less, then the model is considered to have a good fit. The statistical significance of the indirect effects was tested by examining bootstrapped (*k* = 5000) 95% confidence intervals (CIs).

The direct and indirect pathways between depressive symptoms and HOMA2-IR were tested in two stages:

#### Stage 1

We first assessed a cross-lagged panel model to test the temporal ordering of depressive symptoms, sleep-wake disturbance, and BMI (see Figure 1). A key assumption of our model is that depressive symptoms precede BMI and sleep-wake disturbance. We also included autoregressive paths (e.g. the relationship between depressive symptoms at 21 and 30 years) to model stability of these variables and account for confounding effects of this stability on other relationships (e.g. if there is a relationship between depressive symptoms and BMI, changes in depressive symptoms may be attributed to BMI, rather than illness progression). Given that HOMA2-IR was not available at 21 years we were unable to test the direction of its relationship to other variables.

**Figure 1.**
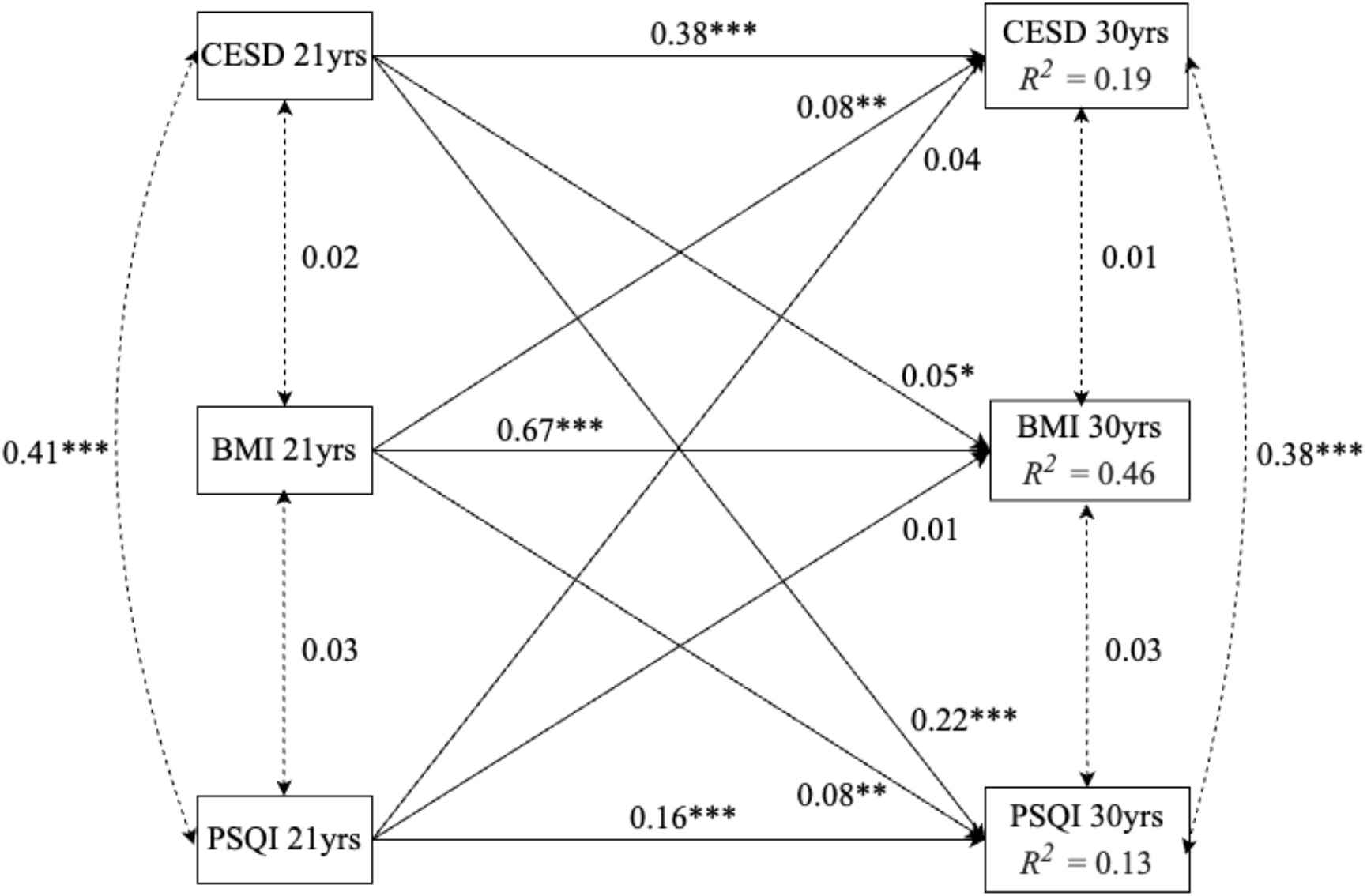
Model 1: Cross lagged panel analysis of longitudinal relationships between depression, sleep, and BMI ***Note***. \*\*\**p* < 0.001; \*\**p* < 0.01; \**p* < 0.05

#### Stage 2

Path analysis was then used to test the hypothesized relationships between depressive symptoms, sleep, BMI, and HOMA2-IR as shown in Figure 2. We used a parallel mediation model that allowed us to compare the relative strength of three competing pathways. As in Stage 1, we included autoregressive relationships, to account for stability of variables over time, we also controlled for cross-sectional relationships between key mediators.

**Figure 2.**
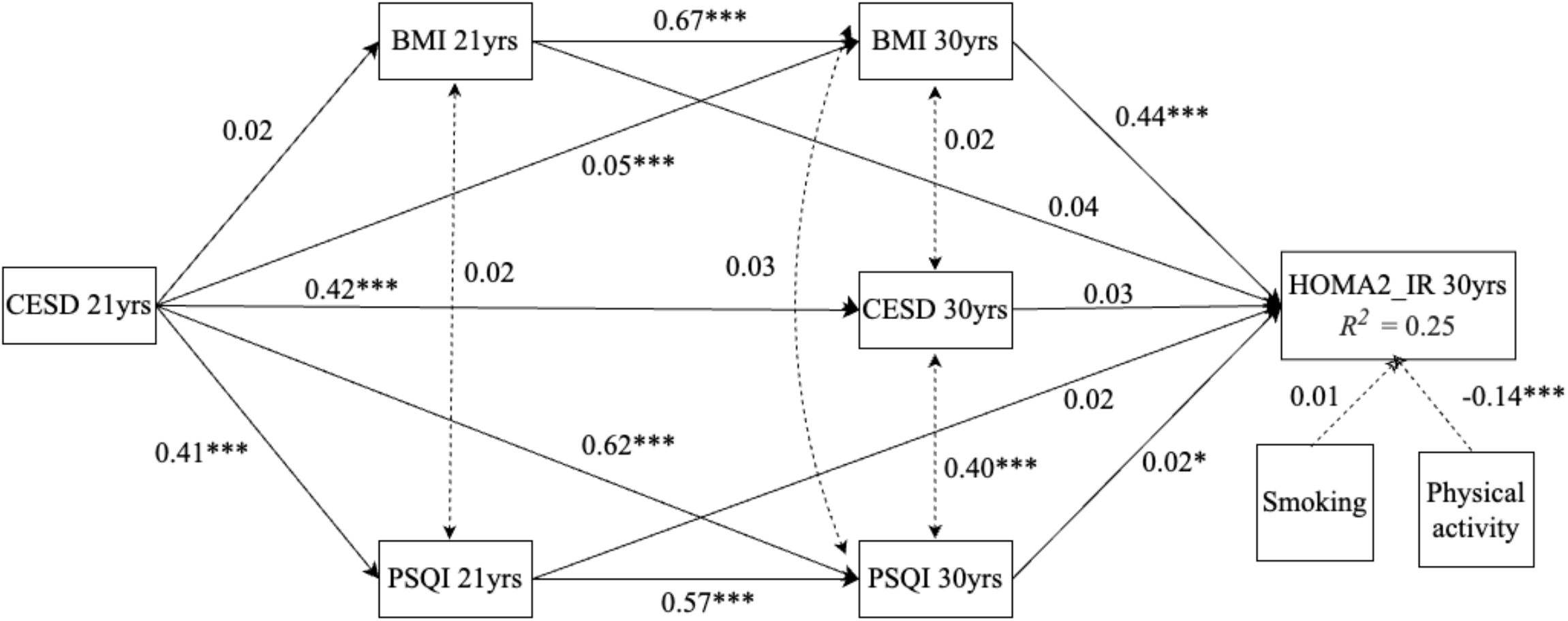
Model of direct and indirect (through BMI and sleep-wake disturbance) relationships between depression and HOMA2_IR. ***Note***. \*\*\**p* < 0.001; \*\**p* < 0.01; \**p* < 0.05

## RESULTS

### Bivariate relationships between depressive symptoms, sleep, and metabolic variables

Correlation analysis found that there were significant cross-sectional and longitudinal relationships between depressive symptoms, sleep-wake disturbance, and cardiometabolic variables. As shown in Table 2, there was a large^28^ significant relationship between depressive symptoms at 21 years and sleep-wake disturbance at 21 years (*r*_s_ = 0.39, p <0.001) but not BMI. BMI and sleep-wake disturbance were not cross-sectionally related at 21 years. At 30 years, there was a small relationship between depressive symptoms and HOMA2-IR (*r*_s_ = 0.07, p <0.01) and BMI (*r*_s_ = 0.05, p <0.05). sleep-wake disturbance (*r*_s_ = 0.29, p <0.001). There were also significant cross-sectional relationships between HOMA2-IR at age 30 and both sleep-wake disturbance (*r*_s_ = 0.13, p <0.001), and BMI (*r*_s_ = 0.48, p <0.001).

**Table 2.** Pairwise cross-sectional and longitudinal correlations (Pearsons’ correlation coefficient) between depression, sleep-wake disturbance, and metabolic variables, controlling for age and gender.

|  | CESD 21yrs | PSQI 21yrs | BMI 21yrs | CESD 30yrs | PSQI 30yrs | BMI 30yrs |
| --- | --- | --- | --- | --- | --- | --- |
| PSQI 21yrs | 0.39*** |  |  |  |  |  |
| BMI 21yrs | 0.01 | 0.02 |  |  |  |  |
| CESD 30yrs | 0.40*** | 0.18*** | 0.05 |  |  |  |
| PSQI 30yrs | 0.29*** | 0.25*** | 0.08** | 0.49*** |  |  |
| BMI 30yrs | 0.05* | 0.03** | 0.66*** | 0.06* | 0.10*** |  |
| HOMA2_IR 30yrs | 0.07** | 0.07* | 0.33*** | 0.10*** | 0.13*** | 0.48*** |

*Note.* \*\*\* $p < 0.001$ ; \*\* $p < 0.01$ ; \* $p < 0.05$

Longitudinally, HOMA2-IR at age 30 was significantly related to depressive symptoms (*r*_s_ = 0.07, p <0.01), sleep-wake disturbance (*r*_s_ = 0.07, p <0.05), and BMI (*r*_s_ = 0.33, p <0.001) at age 21. Depressive symptoms at age 21 was also related to depressive symptoms (*r*_s_ = 0.40, p <0.001), sleep-wake disturbance (*r*_s_ = 0.29, p <0.001), and BMI at age 30 (*r*_s_ = 0.05, p <0.001). Sleep-wake disturbance at age 21 was related to sleep-wake disturbance (*r*_s_ = 0.25, p <0.001), depressive symptoms (*r*_s_ = 0.18, p <0.001), and BMI (*r*_s_ = 0.03, p <0.01) at age 30 (*r*_s_ = 0.05, p <0.05). Meanwhile, BMI at age 21 was significantly related to BMI (*r*_s_ = 0.66, p <0.001) and sleep-wake disturbance (*r*_s_ = 0.08, p <0.01) at age 30, but not to depressive symptoms.

### Stage 1: Cross-lagged effects

We used a cross-lagged panel model to test the direction of the relationship between depressive symptoms, sleep-wake disturbance and BMI (given that this was the only metabolic outcome available at both time points) whilst controlling for age, gender, smoking, and physical activity at 30-years (Figure 1). The chi-square was significant indicating poor model fit (χ^2^ = 92.75, p < 0.001), however other indices suggested the path model adequately fit with the data; RMSEA = 0.06 [0.05, 0.07]; SRMR. = 0.04; CFI = 0.97; TLI = 0.89. Regression coefficients suggested that depressive symptoms at 21 years predicted depressive symptoms at 30 years (see Figure 1 and Table 3; *B =* 0.38, *p* <0.001), BMI at 21 years was uniquely associated with BMI at 30 years (*B =* 0.67, *p* <0.001), and sleep-wake disturbance at 21 years was related to sleep-wake disturbance at 30 years (*B =* 0.16, *p* <0.001).

**Table 3.** Cross-lagged effects and ANOVA of differences regarding longitudinal pathways from depression, BMI and sleep.

| **Relationship** | ***B (SE)*** | **CI 95** |
| --- | --- | --- |
| CESD 21 years → CESD 30 years | 0.38*** (0.03) | [0.32, 0.45] |
| BMI 21 years → BMI 30 years | 0.67*** (0.02) | [0.63, 0.71] |
| PSQI 21 years → PSQI 30 years | 0.16*** (0.03) | [0.11, 0.21] |
| CESD 21 years → BMI 30 years | 0.05* (0.02) | [0.01, 0.09] |
| BMI 21 years → CESD 30 years | 0.08** (0.02) | [0.03, 0.13] |
| *χ<sup>2</sup> of differences* | 1.11 |  |
| PSQI 21 years → BMI 30 years | 0.01 (0.02) | [-0.03, 0.05] |
| BMI 21 years → PSQI 30 years | 0.08** (0.02) | [0.03, 0.13] |
| *χ<sup>2</sup> of differences* | 4.85* |  |
| CESD 21 years → PSQI 30 years | 0.22*** (0.03) | [0.17, 0.28] |
| PSQI 21 years → CESD 30 years | 0.04 (0.03) | [-0.01, 0.09] |
| *χ<sup>2</sup> of differences* | 26.72*** |  |
| ***Co-variances*** |  |  |
| CESD 21 yrs + BMI 21 yrs | 0.02 (0.03) | [-0.03, 0.07] |
| PSQI 21 yrs + BMI 21 yrs | 0.03 (0.03) | [-0.02, 0.09] |
| CESD 21 yrs + PSQI 21 yrs | 0.41*** (0.03) | [0.35, 0.48] |
| CESD 30 yrs + BMI 30 yrs | 0.01 (0.02) | [-0.02, 0.05] |
| PSQI 30 yrs + BMI 30 yrs | 0.03 (0.02) | [-0.00, 0.07] |
| CESD 30 yrs + PSQI 30 yrs | 0.38*** (0.03) | [0.32, 0.44] |

**Note.** \*\*\* $p < 0.001$ ; \*\* $p < 0.01$ ; \* $p < 0.05$

We found the cross-lagged relationship between depressive symptoms and BMI was bidirectional. Depressive symptoms at 21 years was uniquely associated with BMI at 30 years (*B =* 0.05, *p* = 0.025) and the reverse was also true (*B =* 0.08, *p* = 0.001). An analysis of variance (see Table 3; ANOVA) showed that there was not a significant difference between effect sizes (*χ*^*2*^ = 1.11, *p = 0*.*21*) suggesting that neither BMI nor depressive symptoms was more strongly predictive of the other. Meanwhile, depressive symptoms at 21 years was uniquely associated with sleep-wake disturbance at 30 years (*B =* 0.22, *p* <0.001) but sleep-wake disturbance at 21 years was not related to depressive symptoms at 30 years (*B =* 0.04, *p =* 0.10). ANOVA confirmed that there was a significant difference between these relationships (χ^2^ *=* 26.72, *p* <*0*.*001)*, indicating that depressive symptoms is more predictive of sleep-wake disturbance than the reverse. Conversely, BMI at 21 years led to sleep-wake disturbance at 30 years (*B =* 0.08, *p* = 0.001), not the other way around (*B =* 0.01, *p* = 0.50). ANOVA showed that the difference between these effect sizes was significant (χ^2^ = 4.85, p <0.05), confirming that BMI is a stronger predictor of sleep-wake disturbance than the reverse.

### Stage 2: Direct and indirect relationships between depressive symptoms and HOMA2-IR

We also hypothesized that both BMI and sleep-wake disturbance would mediate the relationship between depressive symptoms and HOMA2-IR. As shown in Figure 2 and Table 4 we tested three potential indirect pathways to metabolic dysfunction for individuals with heightened depressive symptoms, to assess their relative strength; path a was through BMI at 21 and 30 years, path b was through sleep disturbance at 21 and 30 years, and path c was though depressive symptoms at 30 years. The chi-square was significant indicating poor model fit (χ^2^ = 50.848, p < 0.001), however other indices suggested the path model adequately fit with the data; RMSEA = 0.05 [0.04, 0.07]; SRMR. = 0.03; CFI = 0.98; TLI = 0.92.

**Table 4.** Standardized regression coefficients showing direct and indirect relationships between depression at 21 years and HOMA2_IR at 30 years.

| **Direct and indirect pathways** | **Estimate (SD)** | **CI 95** |
| --- | --- | --- |
| CESD 21yrs → Sleep 21yrs → HOMA2_IR 30yrs | 0.00 (0.01) | [-0.02, 0.03] |
| CESD 21yrs → Sleep 30yrs → HOMA2_IR 30yrs | 0.01* (0.00) | [0.00, 0.02] |
| a. CESD 21yrs → Sleep 21yrs → Sleep 30yrs → HOMA2_IR 30yrs | 0.01* (0.00) | [0.00, 0.01] |
| CESD 21yrs → BMI 21yrs → HOMA2_IR 30yrs | 0.00 (0.00) | [0.00, 0.00] |
| CESD 21yrs → BMI 30yrs → HOMA2_IR 30yrs | 0.02* (0.01) | [0.01, 0.04] |
| b. CESD 21yrs → BMI 21yrs → BMI 30yrs → HOMA2_IR 30yrs | 0.01 (0.01) | [-0.01, 0.02] |
| c. CESD 21yrs → CESD 30yrs → HOMA2_IR 30yrs | 0.01 (0.01) | [-0.01, 0.03] |
| Contrast 1: a - b | -0.00 (0.01) | [-0.01, 0.03] |
| Contrast 2: a - c | -0.00 (0.01) | [-0.03, 0.02] |
| Contrast 3: b - c | -0.00 (0.01) | [-0.03, 0.02] |
| Total indirect | 0.01 (0.01) | [-0.01, 0.04] |
| Direct relationship: CESD 21yrs → HOMA2_IR 30yrs | 0.04 (0.03) | [-0.02, 0.10] |
| **Total relationship** | **0.05\* (0.03)** | **[0.00, 0.10]** |

*Note.* \*\*\* $p < 0.001$ ; \*\* $p < 0.01$ ; \* $p < 0.05$

As shown in Figure 2, regression coefficients showed that both BMI (*B =* 0.22, *p* <0.001) and sleep-wake disturbance (*B =* 0.22, *p* <0.001) at 30 years were uniquely associated with HOMA2-IR at 30 years, whereas depressive symptoms at 30 years was not (*B =* 0.03, *p =* 0.34). Neither BMI (*B =* 0.04, *p* = 0.18) nor sleep-wake disturbance (*B =* 0.02, *p* = 0.24) at 21 years were related to HOMA2-IR at 30 years.

Despite earlier finding a significant bivariate correlation between depressive symptoms at 21 years and HOMA2-IR at 30 years, when we looked at a more complex model there was no direct relationship between these variables (*B =* 0.04, *p* = 0.24). Sleep-wake disturbance (see Table 4; *B =* 0.01, *p* = 0.018) and BMI at 30 years (*B =* 0.02, *p* = 0.006) were both found to be significant indirect pathways, however once we accounted for autocorrelations, only sleep-disturbance remained a significant indirect pathway (*B =* 0.01, *p* = 0.031), whereas BMI was no (*B =* 0.005, *p* = 0.50). Even so, contrast analysis showed that the strength of indirect effects through sleep-wake disturbance (at 21 and 30 years), BMI (at 21 and 30 years) and depressive symptoms (at 30 years) were not significantly different, meaning no pathway was more influential on HOMA2-IR at 30 years. Taken together, the total relationship between depressive symptoms at 21 years and HOMA2-IR at 30 years was found to be significant (*B =* 0.05, *p* = 0.039) despite no direct relationship.

## DISCUSSION

Our results challenge conventional wisdom that weight gain is the primary driver of metabolic dysfunction for those with major mood disorders. BMI did not significantly mediate this relationship after accounting for autoregressive effect, likely because depressive symptoms and BMI were not cross-sectionally related at 21 years. Thus, weight gain does not fully explain the relationship between depressive symptoms and HOMA2-IR, particularly during young adulthood. Obesity is strongly related to insulin resistance, and accounts for approximately 25% of the variability of insulin sensitivity.^29–31^ However, only one third of individuals who are obese develop insulin resistance.^29–31^ Likewise, our own research has shown that youth accessing early intervention mental health services show elevated HOMA2-IR levels before increased rates of being overweight and obese. Having said this, there was a strong path from BMI at 21 years and 30 years to HOMA2-IR at 30 years suggesting weight loss may still be a useful target for intervention.^13,14,32^ These results have important clinical implications. Weight gain is a common reason for discontinuing effective psychotropic medications however may not explain the increased rates of metabolic syndrome and CVD in individuals with major mood disorders.^33,34^ Simultaneously, existing cardiometabolic guidelines should be updated, so that HOMA2-IR is screened alongside BMI in early intervention services.^35^

Our longitudinal study also tested the hypothesis that sleep-wake disturbance is a key mediator of abnormal glucose-insulin homeostasis in individuals with depressive symptoms. Our findings partially support this hypothesis—while we did identify a significant indirect pathway through sleep-wake disturbance linking depressive symptoms at age 21 to HOMA2-IR at age 30, the effect size was notably small, suggesting that sleep-wake disturbance plays a modest rather than major role in this relationship. Previous research with the MUSP cohort has shown that sleep difficulties at age 2-4 are associated with poorer health quality in adolescence and young adulthood.^36,37^ Sleep-wake interventions such as melatonin and CBT-I have also been shown to improve HOMA2-IR in youth with mood disorders.^16–18^ These results have important implications for future research. Longitudinal research is needed in youth cohorts with mood disorders given these populations often present with early indicators of metabolic dysfunction despite having limited exposure to risk factors such as medication and smoking.^35^

These contributions notwithstanding, the current study has significant limitations that should be considered. Fasting insulin and glucose were not assessed at 21 years meaning we were not able to look at the direction of the relationship between depressive symptoms and metabolic blood markers, particularly HOMA2-IR, that are more sensitive indicators of metabolic disturbance.^12,13^ Additionally, the PSQI is a self-report assessment of sleep quality. Previous research has found that objective measures of sleep-wake activity such as wrist-worn accelerometers or actigraphy devices are more predictive of metabolic outcomes such as adiposity markers, blood lipids, and insulin sensitivity, over and above self-report measures of sleep-wake disturbance.^38,39^ These limitations only heighten the need for more longitudinal tracking of sleep-wake disruption and metabolic dysfunction in youth populations using more robust measures to establish the true size of the relationship.

Moreover, the current research was conducted with a general population cohort and did not compare relationships between subgroups based on mood disorder diagnosis. Effect sizes of indirect pathways were close to 0, suggesting only a small influence of depressive symptoms on HOMA2-IR. However, both sleep-wake disturbances and metabolic dysfunction occur in a subset of individuals with major mood disorders, potentially indicating the existence of syndromes with unique pathophysiological illness pathways.^10,40,41^ It is therefore important for future research to replicate these analyses with clinical populations in early interventions settings.

Overall, the current study challenges common assumptions that weight gain is the primary diver of metabolic dysfunction for those with major mood disorders. Whilst there was a strong path from BMI at 21 and 30 years to HOMA2-IR at 30 years, BMI did not mediate the relationship between depressive symptoms and HOMA2_IR, suggesting other processes are at play. Sleep-wake disturbance was a significant mediator; however indirect pathways were small when looking at a general population. There is an ongoing need for more detailed monitoring and targeted research in clinical youth populations, to establish key drivers of metabolic dysfunction in those with major mood disorders and develop effective early interventions.

## Data Availability

All data produced in the present study are available upon reasonable request to the authors

## DATA AVAILABILITY STATEMENT

Data from the Mater-University of Queensland Study of Pregnancy may be available on request.

## Notes

### Competing Interest Statement

EMS COI:
Associate Professor Elizabeth Scott is a Principal Research Fellow at the Brain and Mind Centre University of Sydney a Consultant Psychiatrist and Adjunct Clinical Professor at the School of Medicine University of Notre Dame. She previously served as the Discipline Leader for Adult Mental Health at Notre Dame until January 2025. In addition she is a member of Medibanks Medical and Mental Health Reference Groups. A/Prof Scott has also delivered educational seminars on the clinical management of depressive disorders receiving honoraria from pharmaceutical companies including Servier Janssen and Eli Lilly. Moreover she has contributed to a national advisory board for Pfizers antidepressant Pristiq and served as the National Coordinator for an antidepressant trial sponsored by Servier
IBH COI:
Professor Hickie is a Professor of Psychiatry and the Co-Director of Health and Policy Brain and Mind Centre University of Sydney. He has led major public health and health service development in Australia particularly focusing on early intervention for young people with depression suicidal thoughts and behaviours and complex mood disorders. He is active in the development through codesign implementation and continuous evaluation of new health information and personal monitoring technologies to drive highly-personalised and measurement-based care. He holds a 3.2% equity share in Innowell Pty Ltd that is focused on digital transformation of mental health services

### Funding Statement

JJC is supported by a NHMRC Emerging Leadership Fellowship (2008196); IBH is supported by a NHMRC Leadership L3 Fellowship (2016346); FI is funded by a NHMRC Emerging Leadership Fellowship (2018157). SM is supported by the Cottle Family Fellowship in Youth Mental Health. WC and MKC were supported by the Australian Government Research Training Program (RTP) Scholarship.

### Author Declarations

The University of Queensland Human Research Ethics Committee (mothers: B/555/SS/01/NHMRC - 30/11/2001, offspring: B/660/SS/01/NHMRC - 20/12/2001) and The Mater Human Research Ethics Committee (mothers: 505A - 29/6/2002, offspring: 506A - 15/07/2002).

